# COVID-19 prognostic model using Bayesian networks learnt on patient data

**DOI:** 10.1101/2022.10.24.22281436

**Authors:** Nikolas Bernaola, Concha Bielza, Pedro Larrañaga

**Affiliations:** Computational Intelligence Group. Departamento de Inteligencia Artificial. Universidad Politécnica de Madrid, Spain

## Abstract

The response to the ongoing second wave of the COVID-19 pandemic can be helped by giving medical professionals access to models learned on patient data. To achieve this, we learned a Bayesian network model to predict risk of ICU admission, death and time of stay in the hospital from patient history, initial vital signs, initial laboratory tests and medication. Data were obtained from patients that were admitted to an HM hospital with suspicion of COVID-19 until 24/04/2020, excluding unconfirmed diagnosis, those who were admitted before the epidemic started in Madrid, had an outcome that was not discharge or death or died within 24 hours of presentation. Relevant variables for the model were selected with help from medical professionals. We learned the model using Bayesian search as implemented in GeNIe. Of 2,307 patients in the dataset, 679 were excluded. With the remaining 1,645 patients, we learned a model that predicted death with 86.4% accuracy. Some of the initial variables were discarded because they were independent of the outcomes of interest conditioned on some of the other variables. This high redundancy might be useful to build simpler tests for the severity of COVID-19. We show how the model can be used at different stages of patient admission and even with only partial information about the patient. This can be done by clinicians that want a fast second opinion or a summary of the available data from previous patients similar to the one at hand. We then include how we plan to improve the model with extra patient data and how it could be expanded to other contexts, like for example, an epidemiological one.

## Introduction

The coronavirus disease 2019 (COVID-19) pandemic, declared by the WHO Director General at the media briefing on March 11^th^ 2020, is caused by the named Severe Acute Respiratory Syndrome Coronavirus 2 (SARS-CoV-2)^1^. It started in Wuhan, China, but it later spread to the rest of the world, with as of the last week of 2020, over 84.53^2^ million confirmed cases worldwide.

Spain, with 1,958,844 confirmed cases and 51,078 confirmed deaths as of December 31th 2020^2^, has been undergoing a very strong second wave (see Fig. 1) of the pandemic over the fall with a peak in November and cases on the rise again with 297 cases notified over the last 14 days per 100,000 inhabitants^2^.

**Figure 1:**
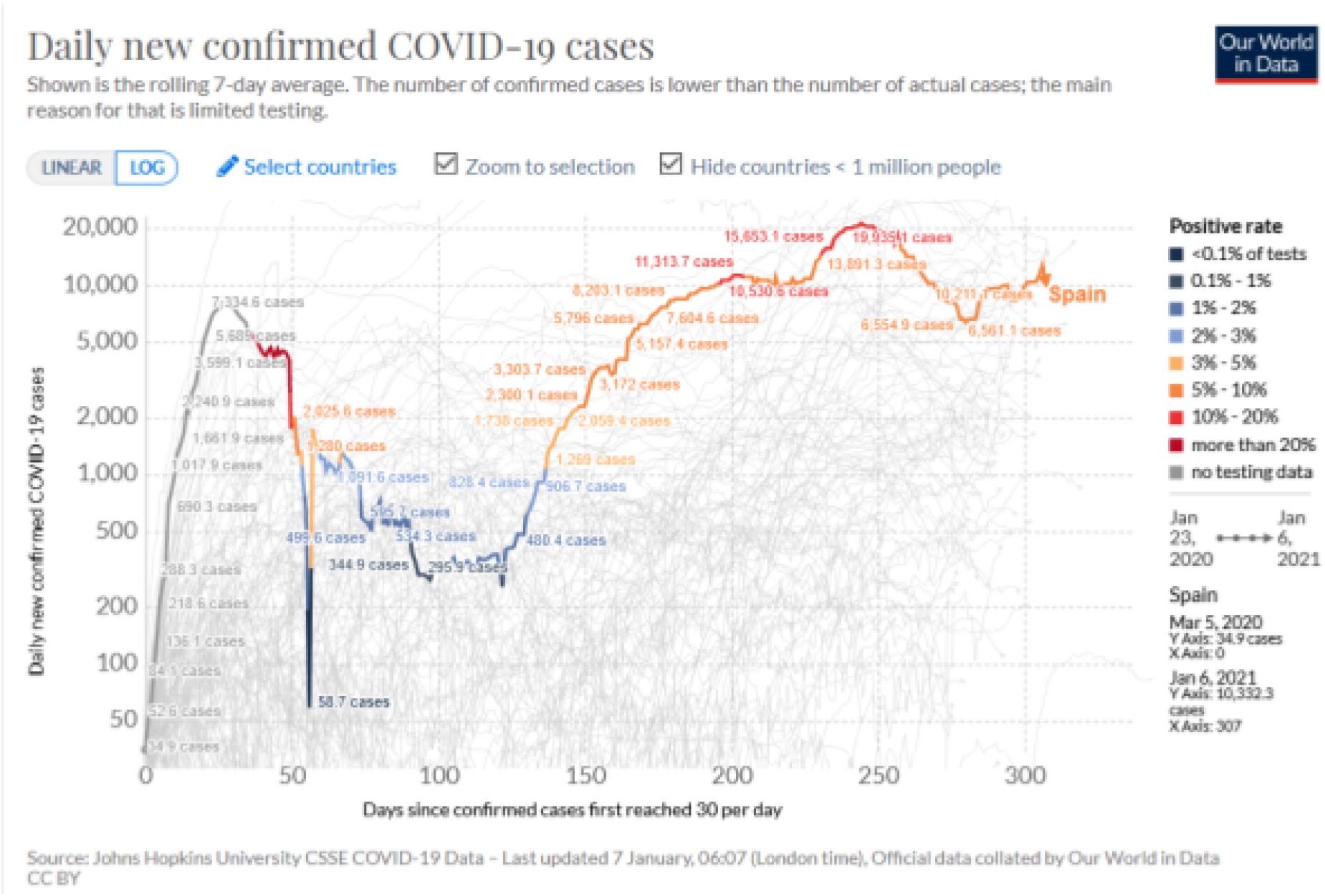
Daily new confirmed cases of COVID-19 in Spain. It clearly shows the ongoing second wave, very high test positivity rate and how after a peak in November, the cases are slightly on the rise again. (Data by Johns Hopkins University, chart by our World in Data^9^)

Because it is a new epidemic, the specific mechanisms and pathophysiology remain elusive, with clinical experience showing significant heterogeneity in the development of symptoms in severe acute cases^3^. The analysis of data on the clinical characteristics, received treatment and outcomes of COVID-19 patients is of vital importance to reduce its mortality, target treatment to presentation of the disease and help with triage and proper management of hospital resources. There has been previous work on finding prognostic models and clinical predictors for COVID-19^4,5,6,7^ but it has been found to be flawed^8^ due in part to poor reporting, no explanation of the intended use of the models and no description of the study population. There is also a lack of work of this type in Spain with, to the best of our knowledge, only one other study in collaboration with a hospital in Madrid^7^.

For these reasons, in this article we present a Bayesian network model learnt using data from 1,645 patients admitted with symptoms of COVID-19 to hospitals of the HM network in Madrid during the first wave of the pandemic. This work has been done in partnership with various medical professionals which helped with an initial analysis and reporting of the study population^10^ and further posed concrete questions for the model to answer so that there is a clear use case.

## Methods

### Data sources

We obtained the data from the HM hospital network in Madrid, thanks to its project ‘COVID DATA SAVE LIVES’^11^. This anonymized clinical dataset comes from the HM hospitals HER system. It was openly released on April 25th on demand to research groups that wanted to analyse it, provided they presented a project beforehand and said project was approved by the corresponding board of experts.

The data included patients’ age, gender, past diagnoses, smoking status, admission data, initial vital signs and complementary tests performed in the Emergency Room, vitals and tests performed throughout their hospital stay, treatments received (including previous medications continued and specific treatment for COVID-19), destination at discharge (or death) and diagnoses during their stay.

### Exclusion criteria

Patients admitted before the first cases were declared in Madrid (24/02/2020), who had not yet reached an outcome (discharge or death) by 24/04/2020, transferred to a different hospital for admission, voluntarily discharged, with an unconfirmed diagnosis or that were interned for less than 24 hours were excluded from the analysis. After exclusion there were 1,645 patients remaining. The exclusion process is explained in Figure 2.

**Figure 2:**
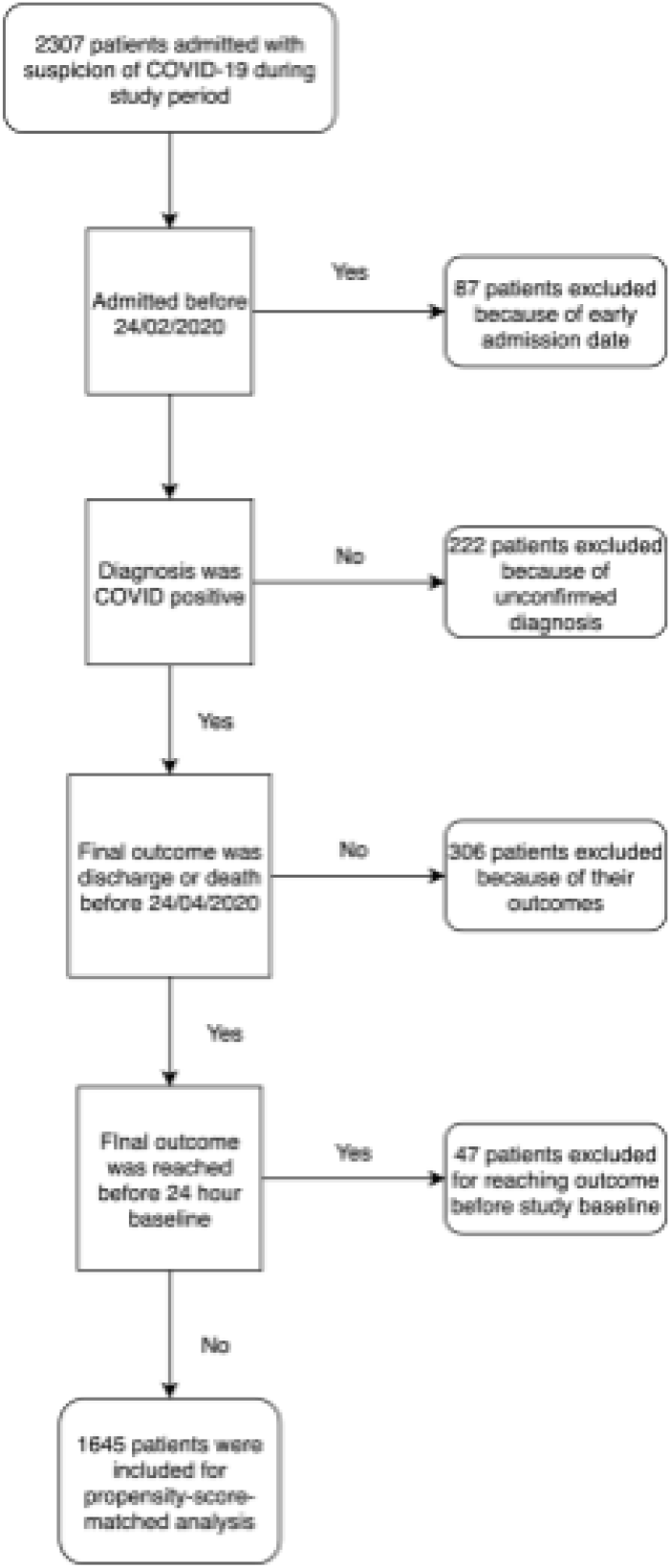
Flow diagram of patient exclusion from the initial sample.

### Model learning

#### Variable selection

Since the dataset had an enormous number of variables for each patient, we could not use all of them for risk of overfitting the model to the comparatively small amount of training data we had.

Therefore, we chose to focus on only a few variables. To do so, we consulted two independent experts on what the most important indicators for prognosis were. These variables are the following: age (divided into the following groups: <40, 40-59, 60-79 and ≥80), gender, past diagnoses (diabetes, hypertension, hypercholesterolemia, ischaemic events [recorded as continued use of anticoagulants and/or aspirin], chronic obstructive pulmonary disease [COPD] and cancer), toxic habits (current or past smoking), allergies (to penicillin and other medications), initial vital signs (temperature, heart rate, oxygen saturation [measured through pulse oximetry] and blood pressure [systolic and diastolic]) and initial laboratory parameters (lactate dehydrogenase [LDH], D-dimer, ferritin, C-reactive protein [CRP], lactate, creatinine, procalcitonine, lymphocyte count and neutrophil count). For these features we took the first analysis they had after arriving at the hospital or, when possible, the average of the first two if taken within 24 hours so as to reduce the amount of missing data. Viral load and Interleukin-6 level were considered too. However, viral load was not available in the data and only 30 patients had been tested for IL-6 level in their first two tests, so those variables were discarded.

We also added sixteen medications that were considered the most important by the experts as binary variables indicating whether a patient had been administered the drug or not. The drugs considered for the network were: four corticoids (methylprednisolone, dexamethasone, hydrocortisone and prednisone); two antivirals (ritonavir with lopinavir, and oseltamivir); seven antibiotics (amoxicillin, piperacillin, linezolid, azithromycin, ceftriaxone, meropenem and levofloxacin); and three immunomodulators (hydroxychloroquine, tocilizumab, and interferon beta 1-b).

Finally, three possible target variables were added: time of stay in the hospital, ICU admission and death, all relevant for the main objective of predicting the severity of the disease in a given patient. Table 1 shows all the predictor variables used for the model. For a complete analysis of the dataset, see previous work by our team^10^. We studied the association of the variables to mortality and used propensity-score matching to find the effect of treatments on patient outcomes.

**Table 1:** Variables considered for the model. In red, the variables that were rejected due to high redundancy. (They were independent of the targets given the rest of the variables in the same group)

| Variable selection |  |  |  |  |
| --- | --- | --- | --- | --- |
| Demographic | Initial vital signs | Initial laboratory parameters | Past diagnoses | Medication |
| Age<br>Gender | Temperature<br>Heart rate<br>Oxygen saturation<br>Blood pressure | LDH<br>D-dimer<br>Ferritin<br>C-reactive protein<br>Lactate<br>Creatinine<br>Procalcitonine<br>Lymphocyte count<br>Neutrophil count | Diabetes<br>Hypertension<br>Hypercholesterolemia<br>Continued use of aspirin<br>Continued use of anticoagulants<br>COPD<br>Cancer<br>Current smoking<br>Past smoking<br>Penicillin allergy<br>Allergy to other medication | Methylprednisolone<br>Dexamethasone<br>Hydrocortisone<br>Prednisone<br>Ritonavir and Lopinavir<br>Oseltamivir<br>Amoxicillin<br>Piperacillin<br>Linezolid<br>Azithromycin<br>Ceftriaxone<br>Meropenem<br>Levofloxacin<br>Hydroxychloroquine<br>Tocilizumab<br>Interferon beta 1-b |

All lab tests but LDH, D-Dimer and lymphocyte count were removed from the network in an effort to improve readability by reducing the number of redundant variables. The same happened with initial blood pressure and the other vital signs. This was done because those variables were found in every network learned to be independent of mortality or ICU admission given the other initial variables. This was done through the d-separation graphical criterion^12^.

#### Missing data

All variables except for the initial laboratory tests and initial vital signs were complete. The ones that were not, were imputed using an iterative imputation procedure, that learned a multivariate regression on all the variables except one against that, then used the newly imputed variable to learn a new regression for the other variables^13^. This process was repeated until the imputed dataset was stable.

#### Bayesian networks

A Bayesian network (BN) is a probabilistic graphical model that combines probability and graph theory to efficiently represent the probability distribution of a group of variables { *X*_1_,,*X*_*n*_ } ^12^. BNs model probabilistic conditional dependencies and independencies between the variables in terms of a directed acyclic graph and a series of conditional probability distributions (CPDs). Each of the nodes in the graph represents a variable with the edges representing conditional (in)dependencies between the variables. Each of the CPDs is associated with a variable *X*_*i*_ and gives the probability distribution of that variable conditioned on its parents in the graph, that is, the nodes that have edges directed towards *X*_*i*_, which we denote **Pa**(*X*_*i*_)The formula for the joint probability distribution of the variables given all the CPDs is:

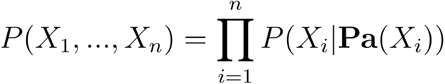

#### Discretization

Continuous variables needed to be discretized, due to limitations of the algorithm. To do so, we followed different strategies depending on the variable in question. For those where it made sense, we consulted the experts to define ranges for which the values were considered normal. This included the initial vital signs and the initial laboratory parameters. We also consulted with them for the age intervals, although given the tremendous age dependency of mortality in COVID-19 cases, more granularity in the upper intervals could be beneficial. Finally, given we didn’t have any clear recommendations for the time in the hospital, we decided to discretize by using equal counts.

#### Bayesian search

Bayesian search of the BN structure is an algorithm introduced by Cooper and Herkovitz in 1992^14^. It is a hill climbing heuristic algorithm guided by a scoring function. In the case of GeNIe’s implementation it is the Bayesian Dirichlet equivalent uniform (BDeu)^15^. This score is optimized by maximizing the probability of the structure of the BN given the dataset used for learning. By Bayes theorem, we have that:

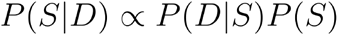

where *P*(*S*|*D*) is the probability of the structure given the data,*P*(*S*) is the prior distribution over the structures and *P*(*D*|*S*) is the probability of the data given the structure (the marginal likelihood). BDeu assumes a uniform prior so that maximizing *P*(*S*|*D*) is the same as maximizing *P*(*D*|*S*) for which there is a closed formula. The assumption of a uniform prior is reasonable since it is mostly uninformative, so no structures are preferred over others initially.

With this score, the algorithm starts with a structure at random and does random edge additions, removals and reversals while checking if the score increases or not. As long as the score increases, the change is accepted. If the score doesn’t change or decreases the change is rejected. If no changes increase the score, the algorithm stops and returns the current network which is the highest scoring one. Then, the algorithm repeats the process with another random starting structure for a number of iterations decided by the user.

Finally, the networks resulting from each iteration are compared and the highest scoring one is returned as the result. The main change in our process was that the comparison between models after each iteration was not done on the basis of the BDeu score but on how well they predicted patient mortality, since it is the most important variable of the three we want to predict.

This accuracy was tested through leave-one-out cross-validation. This was done in an effort to get the best possible model at assessing the severity of each patient.

## Model

The BN model structure is shown in Figure 3. In this state, without any evidence, it serves as a summary of the distribution of the original data. The three variables of interest for prediction are the length of stay, admission to ICU and death. Tables 2a and 2b show accuracy and area under the ROC curve for each of the targets.

**Table 2a:** Accuracy and area under the ROC curve for ICU admission and death target variables.

|  | Death | ICU Admission |
| --- | --- | --- |
| Accuracy | 86.4% | 95.2% |
| AUC | 0.870 | 0.634 |

**Table 2b:** Accuracy and area under the ROC curve for each of the bins of the length of stay target variables.

| Length of stay | Up to 5 days | From 5 to 7 days | From 7 to 11 days | Over 11 days |
| --- | --- | --- | --- | --- |
| Accuracy | 16.3% | 15.8% | 68.6% | 50.1% |
| AUC | 0.702 | 0.642 | 0.592 | 0.745 |

**Figure 3:**
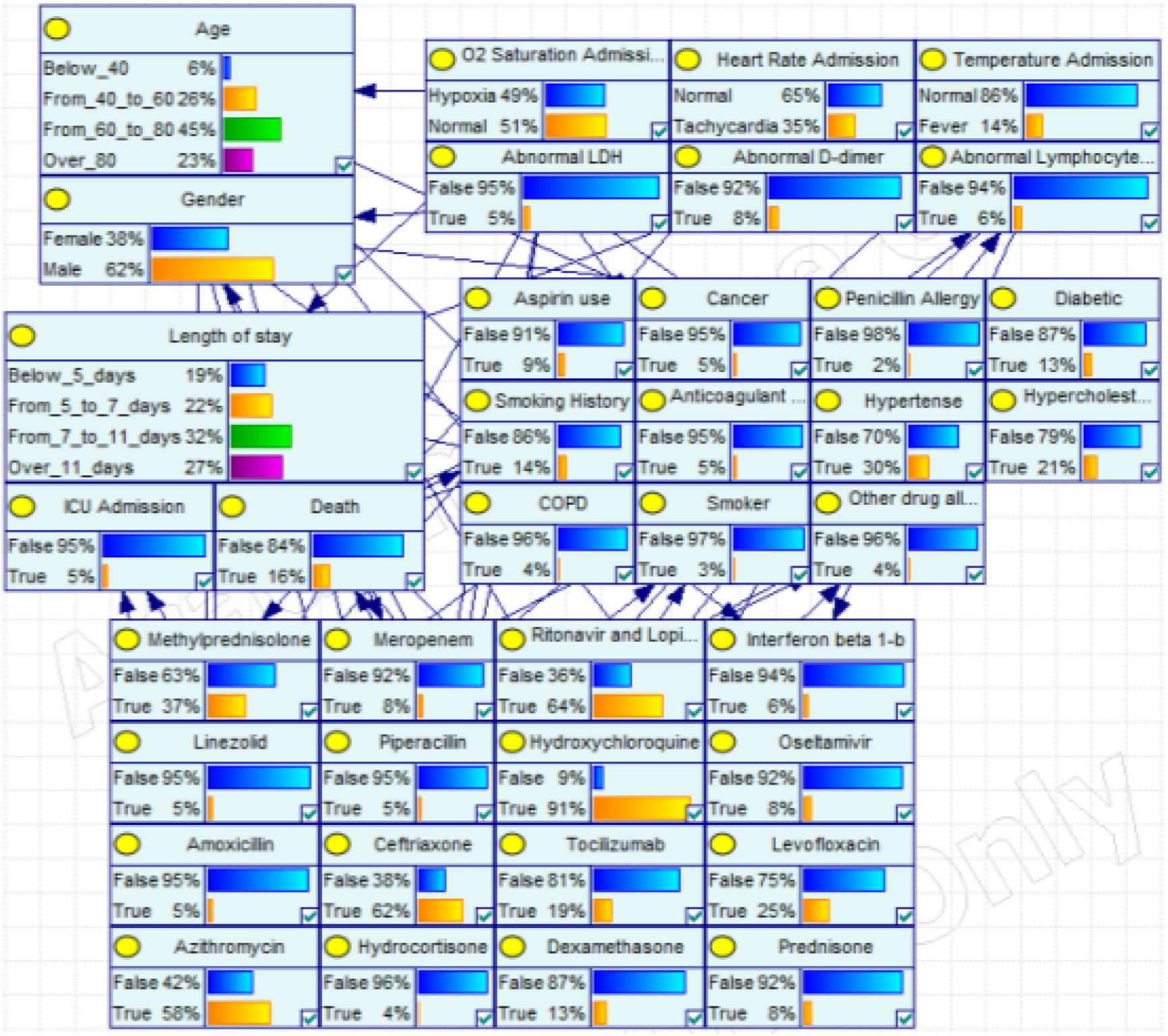
The full Bayesian network model. Without any evidence it serves as a summary of the original data distribution.

Accuracies for the length of stay seem to indicate that the model is generally overestimating the severity of the cases.

### Use case explanation

The model can be used for prognosis at the clinical level (giving doctors the ability to have a quick second opinion that just summarizes the available data), as an example figures 4 to 6 show how the model could be used for a concrete case showing it as we get more information on the severity of the case. From admission (Fig. 4), with only demographic and triage data; followed by adding the initial laboratory tests and past conditions (Fig. 5) and, finally, the medication that the patient is receiving (Fig. 6).

**Figure 4:**
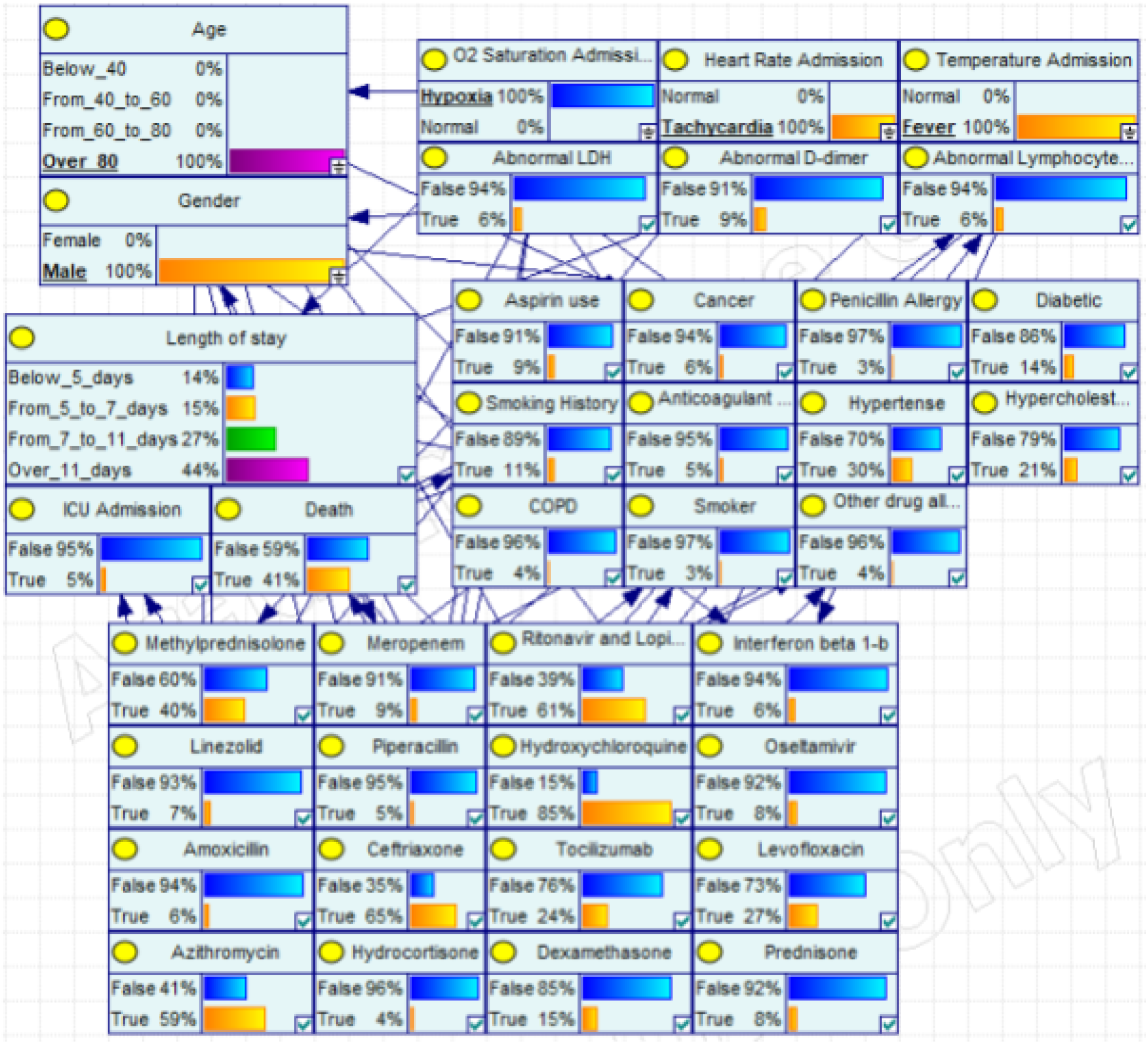
Example at admission. The figure shows an elderly man, over 80 years old, that is admitted with symptoms of COVID-19: hypoxia, fever and increased heart rate. Before we know anything else about the patient, the model already outputs the expected severity of the case (with the probability of death rising to 41%), how long it expects the patient to remain in the hospital and what treatment similar patients have received in the past. Past the clinical level, the probabilities of ICU admission and of length of stay could be used by hospital administrators to be able to make more informed guesses about future occupancy levels and better manage resources.

**Figure 5:**
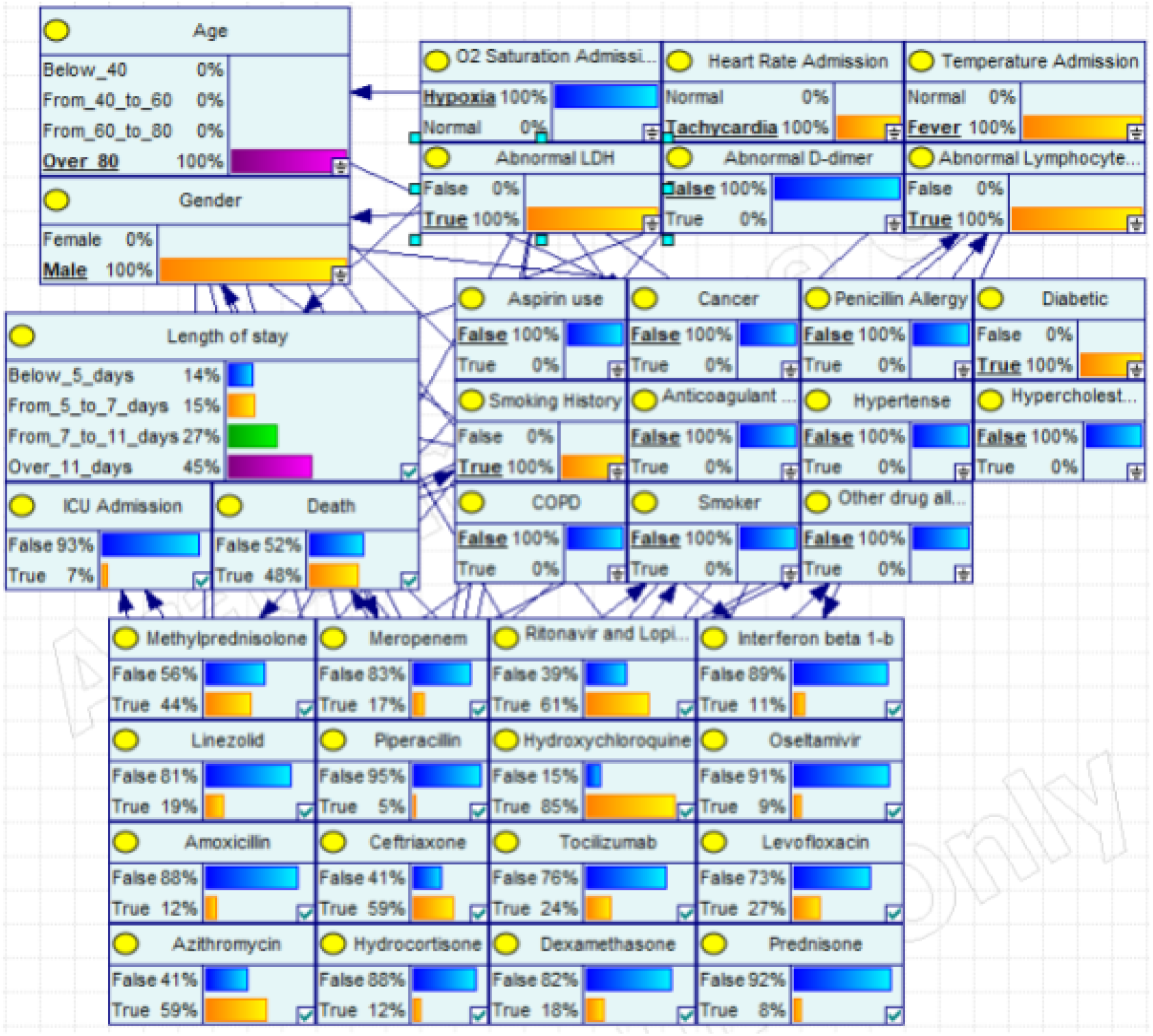
History of the patient and initial tests. In this step, we input the history of the patient, which seems like a controlled diabetic (no hypertension or hypercholesterolemia) that used to smoke. We also add the initial results from the lab, which show a high lymphocyte count and abnormal LDH levels. The evidence is used by the model to update the results, now giving a 48% probability of death and a higher probability of admission to the ICU. This higher risk is also reflected in the medication recommended: probabilities for stronger antibiotics (like meropenem) and steroids (like dexamethasone) have gone up.

**Figure 5:**
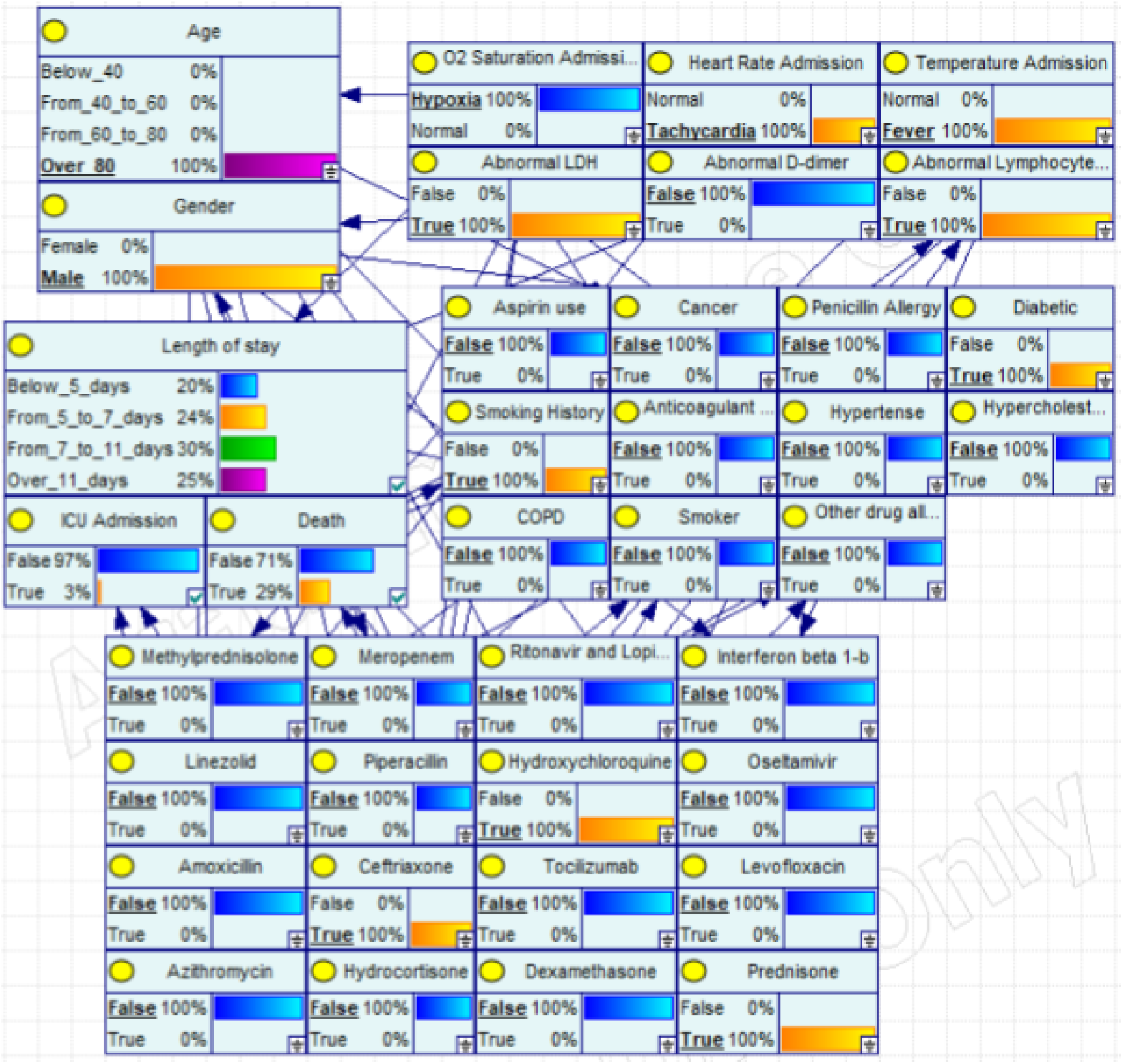
Adding medication. In this final figure, we add the medication that has been given to the patient. In this case, hydroxychloroquine, ceftriaxone and prednisone. This informs the model that the case is less severe than it thought, which updates the estimates of death, ICU admission and length of stay downward. This can be further updated as the case develops, adding new medication, with which the model can keep updating until it resolves.

## Conclusions

We have presented a Bayesian network model learnt using data from patients diagnosed with COVID-19 during the first wave of the pandemic in Spain. Given the current, rapidly worsening, situation we believe that models such as this can be used to help clinicians arrive at conclusions informed by previous patient data much faster which would facilitate the work, especially if the situation worsens and hospital saturation increases.

The model is not a replacement for medical professionals but a tool for informed decision making that will hopefully be useful. Our current goal is to gather as much data as we can so that the model can be improved and then work on building a browser-based app to make it as accessible as possible.

Finally, there is a possible application of this model to an epidemiological setting by taking demographic data and using it to predict incidence of ICU admission and time spent in the hospital if we have an estimate of how much of the population will be infected and how many will need hospitalization. This would help with better resource allocation.

## Data Availability

All data referred to in the manuscript are avaiable upon request to the HM hospitals through the Covid Data saves lives initiative

https://www.hmhospitales.com/coronavirus/covid-data-save-lives/english-version

## Bibliography

1. “Naming the coronavirus disease (COVID-19) and the virus that causes it,” 2020. [Online]. Available: https://www.who.int/emergencies/diseases/novel-coronavirus-2019/technical-guidance/naming-the-coronavirus-disease-(covid-2019)-and-the-virus-that-causes-it [Accessed: 20-Nov-2020]

2. “COVID-19 situation update worldwide, as of 20 October 2020,” European Centre for Disease Prevention and Control, 20-Oct-2020. [Online]. Available: https://www.ecdc.europa.eu/en/geographical-distribution-2019-ncov-cases [Accessed: 20-Nov-2020]

3. Zhou, Fei, et al. “Clinical course and risk factors for mortality of adult inpatients with COVID-19 in Wuhan, China: A retrospective cohort study.” The Lancet (2020).

4. Jianfeng, Xie, et al. “Development and external validation of a prognostic multivariable model on admission for hospitalized patients with covid-19.” medRxiv (2020)

5. Yadaw, Arjun S., et al. “Clinical predictors of COVID-19 mortality.” medRxiv (2020).

6. Soto-Mota, Adrian, et al. “The low-harm score for predicting mortality in patients diagnosed with COVID-19: A multicentric validation study” medRxiv (2020).

7. Torres-Macho, Juan, et al. “The PANDEMYC Score. An easily applicable and interpretable model for predicting mortality associated with COVID-19.” Journal of Clinical Medicine 9.10 (2020): 3066.

8. Wynants, Laure, et al. “Prediction models for diagnosis and prognosis of covid-19: Systematic review and critical appraisal.” BMJ 369 (2020).

9. Max Roser, et al. (2020) - “Coronavirus pandemic (COVID-19)”. [Online] Available: https://ourworldindata.org/coronavirus [Accessed: 20-Nov-2020]

10. Bernaola, Nikolas, et al. “Observational Study of the Efficiency of Treatments in Patients Hospitalized with Covid-19 in Madrid”. medRxiv, 2020. Available: https://www.medrxiv.org/content/10.1101/2020.07.17.20155960v1

11. HM Hospitales, “COVID data save lives” [Online]. Available:https://www.hmhospitales.com/coronavirus/covid-data-save-lives. [Accessed: 20-Nov-2020]

12. Koller, Daphne, and Nir Friedman. Probabilistic Graphical Models: Principles and Techniques. MIT press, 2009.

13. Buck, Samuel F. “A method of estimation of missing values in multivariate data suitable for use with an electronic computer.” Journal of the Royal Statistical Society: Series B (Methodological) 22.2 (1960): 302–306.

14. Cooper, Gregory F., and Edward Herskovits. “A Bayesian method for constructing Bayesian belief networks from databases.” Uncertainty Proceedings 1991. Morgan Kaufmann, 1991, 86–94.

15. Heckerman, David, et al. “Learning Bayesian networks: The combination of knowledge and statistical data.” Machine Learning 20, 3 (1995): 197–243.

